# Distinct promoter regions of the oxytocin receptor gene are hypomethylated in Prader-Willi syndrome and in Prader-Willi syndrome associated psychosis

**DOI:** 10.1101/2021.12.14.21267765

**Authors:** Hannah Marie Heseding, Kirsten Jahn, Christian K. Eberlein, Jelte Wieting, Hannah B. Maier, Phileas Proskynitopoulos, Alexander Glahn, Stefan Bleich, Helge Frieling, Maximilian Deest

## Abstract

**Background:** Prader-Willi syndrome (PWS) is a rare neurodevelopmental disorder caused by a loss of usually paternally expressed, maternally imprinted genes located on chromosome 15q11-q13. Individuals with PWS display a specific behavioral phenotype and have a higher susceptibility than the general population for certain psychiatric conditions, especially psychosis. An impairment of the oxytocin system has been described in Prader-Willi syndrome, but has not yet been investigated on the epigenetic level. Recent studies have pointed out altered methylation patterns of the oxytocin receptor gene (*OXTR*) in various psychiatric disorders, including psychosis.

**Methods:** In this study, we investigated methylation rates of CpG dinucleotides in the promoter region of the oxytocin receptor gene via bisulfite-sequencing using DNA extracted from peripheral blood samples of 31 individuals with PWS and 14 controls matched for age, sex and BMI.

**Results:** Individuals with PWS show significantly lower methylation in the intron 1 region of the *OXTR* than neurotypical controls (p=0.012). Furthermore, male PWS subjects with psychosis show significantly lower methylation of the *OXTR* exon 1 region than those without psychosis (p=0.002). Transcription factor binding site analysis revealed E2F1 as a transcription factor potentially binding to the exon 1 region. E2F1 is physiologically regulated by *Necdin*, an anti-apoptotic protein whose corresponding gene is located within the PWS locus.

**Conclusion:** This study provides evidence of a disruption of the Oxytocin system on an epigenetic level in PWS in general and in individuals with PWS and psychosis.

## Introduction

Prader-Willi Syndrome (PWS) is a rare neurodevelopmental disorder caused by a loss of physiologically paternally expressed genes on chromosome 15q11-q13. In humans, this region is subject to genetic imprinting, resulting in epigenetic silencing of the maternal allele. Thus, only the paternally inherited genes in the 15q11-q13 region are expressed. Loss of these genes in PWS mostly occurs due to either de novo paternal deletion of the 15q11-q13 region (in about 60% of cases, delPWS) or maternal uniparental disomy 15 (about 35% of cases, mUPD) [1]. Less common causes of PWS are defects of the imprinting center due to microdeletions or epimutations and paternal chromosomal rearrangements such as translocations [2].

Predominant clinical features of newborns with PWS include hypotonia and feeding difficulties caused by a poor suck, leading to a failure to thrive [1]. In later infancy, the initial feeding difficulties are replaced by severe hyperphagia, which can lead to life-threatening obesity if food intake is not controlled. Hyperphagia in PWS is most likely due to hypothalamic dysfunction [3]. Other primary features of PWS attributed to disruption of the hypothalamic system include impaired pain perception, sleeping anomalies, temperature instability and multiple endocrine abnormalities such as growth hormone deficiency with short stature, hypogonadism and hypothyroidism. [1, 2, 4]. Nearly all individuals with PWS have a mild to moderate intellectual disability and an overall developmental delay. The distinct behavioral phenotype of PWS is characterized by emotional dysregulation with temper outbursts and impulsivity as well as skin-picking [5]. Furthermore, psychiatric disorders, especially psychosis, anxiety, affective disorders, and autism spectrum disorder (ASD), are more frequent than in the general population [1, 6]. The prevalence of psychiatric characteristics differs between genetic subtypes of PWS. Specifically, PWS individuals with mUPD are at higher risk of developing psychosis after adolescence [7]. Similarly, autism-like behavioral characteristics and diagnosis of ASD is higher in PWS individuals with mUPD than in other PWS subtypes [8–10].

Oxytocin (OT) is a neuropeptide that is produced in the paraventricular (PVN) and supraoptic (SON) nuclei of the hypothalamus. In its function as a neuromodulator, OT is implicated in a complex range of socioemotional cognition and behaviour, such as pair-bond formation, socially reinforced learning and emotional empathy-based behaviours [11]. Furthermore, OT is also involved in regulating body weight, satiety, the reward systems, and the reduction of stress and anxiety [6, 11–13]. In PWS, OT signaling seems to be altered. Abnormal blood levels of OT have been reported in individuals with PWS and a *post mortem* study of five cases has shown a reduction in the volume of the PVN as well as a strong decrease in the number of OT-expressing neurons in the PVN of PWS individuals compared to healthy controls [14–16]. MAGEL2 and NECDIN, two maternally imprinted genes located at the PWS locus, could be linked to dysfunction of OT production. Newborn MAGEL2 knockout mice showed a significant reduction in mature OT in the hypothalamus along with a suckling deficit as seen in human PWS infants, which could be restored by a single postnatal injection of OT [17]. Likewise, a reduction of OT-producing neurons in the hypothalamus was observed in NECDIN deficient mice displaying behavioral characteristics reminiscent of PWS clinical features such as increased skin scraping activity and improved spatial learning and memory [18].

OT mediates its effect by acting on a G-protein coupled receptor (*OXTR*) that is expressed in both brain and peripheral tissues. Single Nucleotide Polymorphisms (SNPs) of the *OXTR* gene have been associated with alterations in sociality and emotional responsiveness in humans and several SNPs could be linked to ASD and schizophrenia [19–21]. Likewise, epigenetic modification of the *OXTR* gene has been studied in regard to a wide range of human socioemotional functioning and psychiatric illnesses [22]. Positive associations have been found between *OXTR* DNA methylation and affect regulation problems and mood deficits, OCD severity, schizophrenia, anorexia nervosa, callous-unemotional traits in youth and social cognitive and communication deficits in ASD [22–28]. In healthy subjects, increased *OXTR* DNA methylation has been associated with decreased functional connectivity between brain regions involved in social perception and decreased functional coupling between the amygdala and emotional regulation areas such as the insular cortex and dorsal anterior cingulate cortex [29]. In ASD individuals, hypermethylation of CpG sites located in the intron 1 of the *OXTR* has been associated with a hypoconnectivity between areas involved in theory of mind (e.g. the superior temporal sulcus) [26]. Moreover, hypermethylation has been linked to prototypic schizophrenic features and both poorer recognition of emotional expressions and smaller volumes in regions associated with social cognition in female schizophrenic patients and controls [30]. Although the oxytocin system is dysregulated in PWS as well as in ASD and psychosis, two major diseases with high prevalence in PWS, no study to date has examined the epigenetic status of the *OXTR* gene in PWS and individuals with PWS and psychosis. We hypothesize that the oxytocin receptor gene is dysregulated in PWS also on an epigenetic level, which could contribute to psychosis in PWS. To investigate our hypothesis, we examine the methylation status of CpG sites located at exon 1 and intron 1 of the *OXTR* gene with a special focus on methylation differences in individuals with PWS and psychosis (figure 1).

**Figure 1:**
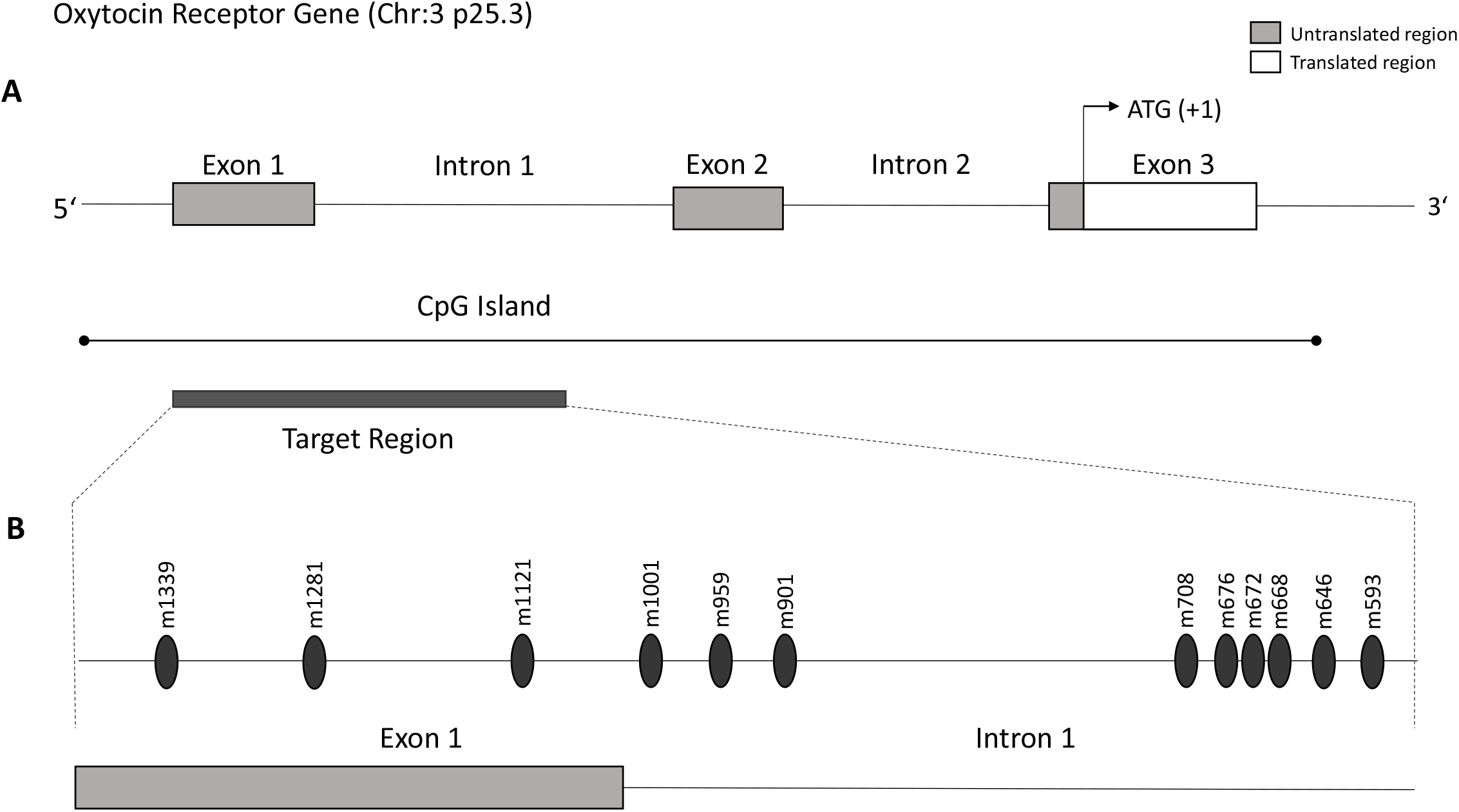
Schematic view of the oxytocin receptor gene. A: The protein-coding (translated) region of the OXTR gene is indicated in white, whereas the untranslated promotor region is indicated in grey. The transcription start site lies within the third exon of the gene, as indicated by the nucleotides ATG. B: The region observed in our study (target region) is located within the first exon and the first intron of the OXTR gene. The position of CpGs is given relative to the transcription start site (example: CpG m1339 is located 1339 nucleotides upstream (5’) of the transcription start site). Only CpGs where significant group differences could be detected are shown in this figure.

## Methods

### Participants

This study adhered to the Declaration of Helsinki and was approved by the local Ethics Committee of Hannover Medical School (Nr. 8129_BO_S_2020). All participants in this study gave their written informed consent for participation after the nature of the procedures had been fully explained. All study participants were recruited at the Outpatient Department for Mental Health in Rare Genetic Disorders of the Department of Psychiatry, Social Psychiatry, and Psychotherapy of Hannover Medical School. Diagnosis of PWS was genetically confirmed. This study cohort is part of a larger registry study entitled PSY-PWS-Germany. PSY-PWS-Germany aims to investigate mental health issues in PWS and is described in more detail elsewhere [31, 32].

EDTA blood samples for the methylation analysis were collected from 32 individuals with PWS and 14 controls. Control subjects had no medical history of psychiatric, neurodevelopmental or neurological conditions and are thus referred to as neurotypical, as opposed to subjects with PWS, whose neurological development differs from the one perceived as “normal” in society [33, 34]. Controls were matched for age (+/-5years), sex and body mass index (BMI). EDTA-blood was stored at Hannover Unified Biobank.

### DNA isolation and bisulfite conversion

Genomic DNA was extracted by the Hannover Unified Biobank from blood using the Hamilton ChemagicStar (Hamilton Germany Robotics, Graefelfing, Germany) and the chemagicStar DNA-Blood1k kit (PerkinElmer chemagen Technology, Baesweiler, Germany).

Bisulfite conversion was performed using the EpiTect 96 Bisulfite Kit (QIAGEN, Hilden, Germany) following the manufacturer’s protocol. Bisulfite treatment converts unmethylated cytosines localized in CpG dinucleotides to uracils (thymines after PCR amplification), whereas methylated cytosines are unaffected by the bisulfite conversion reaction. These changes can later be detected by comparing the sequencing results with the respective reference genomic DNA sequence.

### Primer Design

Primers were manually designed to bisulfite-converted regions of the *OXTR* gene covering the first exon and 490 bases into the first intron using the program Geneious (Biomatters, Auckland, New Zealand). The presence of primer secondary structures was checked with the primer analysis tool Netprimer (https://www.premierbiosoft.com/netprimer/). Melting temperature was checked using the Metabion Biocalculator (http://biotools.nubic.northwestern.edu/OligoCalc.html). The presence of SNPs was checked with the online tool SNPCheck V3 (https://genetools.org/SNPCheck/snpcheck.htm). All primers were ordered from Integrated DNA Technologies (Leuven, Belgium).

### Amplification of the bisulfite-converted target sequences (by touchdown PCRs)

PCR was performed on bisulfite-converted DNA using touchdown PCRs. For the first target, which covers the first exon of the *OXTR* gene, a nested PCR was used to amplify the targeted region. For the second target, which is located in the first intron of the *OXTR* gene, a simple touchdown PCR was performed. For primers and cycling conditions, see supplementary table 1.

All PCRs were performed on a Thermocycler C1000™ (Bio-Rad, Hercules, CA, USA) or a Thermocycler S1000™ (Bio-Rad, Hercules, CA, USA). Amplifying PCRs were performed using the HotStarTaq Master Mix Kit (QIAGEN, Hilden, Germany). Automatic Purification was carried out on a Biomek^®^ NxP using paramagnetic beads (Clean-NGS, GC Biotech^®^, Waddinxveen, The Netherlands). The concentration of purified DNA was measured by Spectrophotometry using the DeNovix DS-11 Fx+ (Denovix, Biozym Scientific GmbH, Hessisch Oldendorf, Germany).

### Sequencing

For sequencing PCR, the BigDye^®^ Terminator v3.1 Sequencing Kit (Applied Biosystems; Foster City, CA, USA) was used. Primers and cycling conditions are listed in the supplementary information. Dye-terminator removal was also automatized on the Biomek^®^ NxP using “Clean DTR” solution (GC Biotech^®^). Sanger sequencing was performed on a HITACHI 3500 XL Genetic Analyser from ABI Life Technologies (Grand Island, NY, USA) according to the manufacturer’s instructions. Taking together the amplified fragments of the Target 1 and Target 2 PCR, a total of 56 CpGs within the first exon and the first intron of the *OXTR* gene were covered (see Fig. 1 for a schematic overview). The position of gene sites is given relative to the transcription start site. The OXTR genetic sequence referred to maps to 8,792,067-8,811,314 in GRCh37 coordinates (from ENSEMBL#ENSG00000180914).

### Analysis of Methylation Rates

Methylation rates were determined using the Epigenetic Sequencing Methylation Software (ESME) software package. ESME aligns the generated sequences and the reference sequence to compare methylation at CpG-sites. For each CpG site per subject, the quantitative methylation information is calculated from the proportion of cytosine and thymine normalized peak values [35].

### Statistical Analysis

Quality of Sequencing was controlled with Sequence Scanner Software 1.0 (ABI Life Technologies). For one sample, the quality value was <20 for Trace Score in the Quality Control Report so that it had to be excluded from further analyses. The Statistical Package for Social Sciences (SPSS, IBM, Armonk, NY) was used for statistical analysis.

Only single CpGs with less than 5% missing values were included, which was the case for all CpGs in our first target. In our second target, one CpG had to be excluded because of more than 5% missing values. Samples with more than 5% missing values were excluded as well, which was the case for one control sample in our first target. In our second target, all samples showed more than 95% valid measurements and could be included. Only CpG sites with greater than or equal to 5% inter-individual variability were included. This applied to 26 of 36 CpG sites in our first target and to all 20 CpG sites of our second target. After applying these criteria for data exclusion, the number of analyzed CpGs was 46.

Mixed linear models were calculated to detect significant group differences in methylation rate. Bonferroni correction method was used to correct for multiple testing. In each analysis, a p-value of <0.05 was considered significant.

### Analysis of Transcription Factor Binding Sites

Transcription factor prediction was performed using the EMBOSS protein database plugin (http://emboss.open-bio.org/) accessing the open-source TransFac database through Geneious 11 (Biomatters, Auckland, New Zealand) with a minimum matching bases of seven for the OXTR gene while allowing for a maximum one base mismatch.

Sequences were cross-referenced using the Factorbook ChipSeq database (https://www.factorbook.org/), thereby also acquiring qualitative information relating to the importance of the CpG nucleotide in question. For transcription factor matching, we selected 8 nucleotides upstream and downstream of each CpG, respectively, and validated the most relevant targets relating to CG importance. For one particular transcription factor, E2F1, the relevance of the CpG nucleotide in question for possible binding was also confirmed through the JASPAR (https://jaspar.genereg.net/) position frequency matrix of this transcription factor.

## Results

### Study Demographics

This study includes 31 individuals with PWS and 14 neurotypical controls, which were matched by age, sex and BMI. Age ranged from 12 to 55 years with a mean age of 28 years. Of the 31 individuals with PWS, 19 were male and 12 were female. In the control group, we included 9 male and 5 female individuals. Distribution of genetic subtypes within the PWS group was deletion in 14 cases, of which 4 patients had the deletion type I, 9 patients had the deletion type II and for one patient, type of deletion was unclear. 9 individuals had the mUPD subtype, 4 had an IC defect and for 4 individuals it was unclear whether they had a mUPD or an IC defect. 7 of the 31 PWS individuals had a history of psychosis, of which 6 were male and one was female. All of the individuals with psychosis had either the mUPD or the IC subtype (table 1).

**Table 1:** Demographic data. Study population is grouped by controls and PWS. PWS individuals are then further grouped by history of psychosis vs no history of psychosis. Abbreviations: Del, deletion; Del I, deletion type I; Del II, deletion tye II, mUPD, maternal uniparental disomy; IC, imprinting defect; ns, not specified because no differentiation of specific subtype could be obtained (Del I vs Del II or UPD vs IC) due to different reasons (e.g. no blood from the parents could be obtained).

Demographics of Study Population
|  |  | Controls | PWS |  |  |
| --- | --- | --- | --- | --- | --- |
|  |  | (n=14) | Total<br>(n=31) | Psychosis<br>(n=7) | No Psychosis<br>(n=25) |
| Age (mean [SD], years) |  | 31.21<br>[8.65] | 27.23<br>[11.63] | 30.57<br>[14.85] | 26.25<br>[10.70] |
| Sex | m | 9<br>(64.3%) | 19<br>(61.3%) | 6<br>(85.7%) | 13<br>(54.2%) |
|  | f | 5<br>(35.7%) | 12<br>(38.7%) | 1<br>(14.3%) | 11<br>(45.8%) |
| BMI (mean [SD], kg/m<sup>2</sup>) |  | 26.62<br>[5.46] | 27.98<br>[7.38] | 31.28<br>[12.16] | 27.02<br>[5.31] |
| Subtype | Del | / | 14 (4 Del I, 9 Del II, 1 ns) | / | 14 (4 Del I, 9 Del II, 1 ns) |
|  | mUPD/IC | / | 17 (9 mUPD, 4 IC, 4 ns) | 7 (2 mUPD, 2 IC, 3 ns) | 10 (7 mUPD, 2 IC, 1 ns) |

### Intron 1 region

As CpG methylation is known to be sex-specific, we first analyzed if there were any differences between males and females. In the intron 1 region, we found no significant effect of sex on the mean methylation rate (F_(1,896)_=3.458; p=0.063).

#### Hypomethylation in PWS subjects compared to neurotypical controls

Next, we analyzed if there were any differences in mean methylation within the intron 1 region between PWS and neurotypical controls. In fact, significant effects were detected for group on the mean methylation rate (F_(1,896)_=6.285; p=0.012). Controls showed a higher mean methylation rate of 31.6% ± 1.3% and PWS individuals showed a lower value of 27.6% ± 0.9%.

Analysis of methylation rates at single CpG sites showed significant effects of group at the following CpG sites: m1001, m959, m901, m708, m676, m672, m668, m646, m593 (table 2A). All of these CpG sites showed hypomethylation in PWS subjects compared to neurotypical controls (figure 2A). It is particularly noteworthy that CpG sites m708, m676, m672, m668, m646 and m593 are located within close proximity of each other and appear to form a cluster where cytosines within CpG sites are methylated less in the PWS group than in controls.

**Table 2.** <u>A: Comparison of methylation rates at single CpG sites in controls and PWS subjects in the intron 1 region.</u> Significant differences in methylation rates between controls and PWS subjects could be detected at CpGs m1001, m959, m901, m708, m676, m672, m668, m646 and m593. <u>B: Comparison of methylation rates at single CpG sites in male PWS subjects with psychosis (yes) and without psychosis (no) in the exon 1 region.</u> Significantly lower methylation rates in subjects with a history of psychosis could be detected at CpG m1339, m1281 and m1121.

| Analyzed CpG | Mean Controls | Standard Error Controls | Mean PWS | Standard Error PWS | Difference in Means | p-value | F-value |
| --- | --- | --- | --- | --- | --- | --- | --- |
| m1001 | 17.1% | 2.0% | 9.9% | 1.4% | 7.3% | 0.004 | 9.020 |
| m989 | 19.5% | 1.8% | 15.5% | 1.2% | 4.0% | 0.071 | 3.439 |
| m982 | 24.7% | 2.0% | 23.0% | 1.3% | 1.7% | 0.500 | 0.462 |
| m959 | 12.5% | 1.3% | 8.7% | 0.9% | 3.8% | 0.023 | 5.592 |
| m934 | 51.4% | 2.3% | 50.8% | 1.5% | 0.6% | 0.823 | 0.051 |
| m924 | 64.4% | 1.8% | 62.2% | 1.2% | 2.2% | 0.315 | 1.035 |
| m901 | 80.8% | 1.3% | 77.5% | 0.9% | 3.2% | 0.050 | 4.069 |
| m860 | 66.6% | 1.7% | 64.3% | 1.2% | 2.4% | 0.260 | 1.302 |
| m835 | 42.2% | 2.5% | 41.1% | 1.7% | 1.1% | 0.709 | 0.141 |
| m826 | 23.8% | 2.0% | 22.2% | 1.3% | 1.6% | 0.514 | 0.433 |
| m808 | 30.8% | 1.7% | 27.1% | 1.1% | 3.7% | 0.079 | 3.232 |
| m806 | 30.4% | 2.2% | 25.9% | 1.5% | 4.5% | 0.100 | 2.821 |
| m774 | 31.8% | 2.1% | 31.8% | 1.4% | 0.0% | 0.996 | 0.000 |
| m719 | 40.9% | 2.2% | 39.0% | 1.5% | 2.0% | 0.462 | 0.550 |
| m708 | 15.2% | 1.8% | 9.1% | 1.2% | 6.1% | 0.007 | 7.925 |
| m676 | 21.1% | 1.9% | 11.2% | 1.2% | 9.9% | < 0.001 | 19.769 |
| m672 | 12.6% | 1.0% | 6.9% | 0.7% | 5.6% | < 0.001 | 22.661 |
| m668 | 10.9% | 1.6% | 6.6% | 1.0% | 4.2% | 0.030 | 5.036 |
| m646 | 11.4% | 1.0% | 4.7% | 0.7% | 6.6% | < 0.001 | 30.563 |
| m593 | 14.7% | 1.1% | 7.9% | 0.7% | 6.8% | < 0.001 | 26.223 |

| Analyzed CpG | Mean Yes | Standard Error Yes | Mean No | Standard Error No | Difference in Means | p-value | F-value |
| --- | --- | --- | --- | --- | --- | --- | --- |
| m1399 | 0.3% | 1.3% | 1.6% | 0.9% | -1.3% | 0.429 | 0.658 |
| m1388 | 2.2% | 0.7% | 2.9% | 0.5% | -0.8% | 0.401 | 0.743 |
| m1383 | 7.0% | 1.9% | 6.6% | 1.3% | 0.4% | 0.868 | 0.028 |
| m1359 | 0.7% | 1.2% | 3.3% | 0.8% | -2.6% | 0.082 | 3.426 |
| m1345 | 3.8% | 1.4% | 5.8% | 1.0% | -1.9% | 0.279 | 1.251 |
| **m1339** | **2.7%** | **1.1%** | **5.7%** | **0.8%** | **-3.0%** | **0.043** | **4.764** |
| m1335 | 1.8% | 1.2% | 3.6% | 0.8% | -1.8% | 0.238 | 1.498 |
| m1312 | 3.3% | 1.1% | 5.5% | 0.8% | -2.1% | 0.138 | 2.421 |
| m1285 | 2.7% | 1.0% | 3.7% | 0.7% | -1.0% | 0.399 | 0.748 |
| m1283 | 6.3% | 1.3% | 8.3% | 0.9% | -2.0% | 0.218 | 1.638 |
| **m1281** | **4.7%** | **1.1%** | **7.5%** | **0.7%** | **-2.8%** | **0.048** | **4.522** |
| m1279 | 7.0% | 1.5% | 9.6% | 1.0% | -2.6% | 0.165 | 2.102 |
| m1258 | 6.2% | 1.6% | 9.1% | 1.1% | -2.9% | 0.159 | 2.175 |
| m1254 | 1.8% | 1.9% | 3.8% | 1.3% | -1.9% | 0.400 | 0.745 |
| m1234 | 5.7% | 1.4% | 8.0% | 0.9% | -2.3% | 0.172 | 2.028 |
| m1219 | 1.8% | 0.6% | 1.7% | 0.4% | 0.1% | 0.850 | 0.037 |
| m1201 | 13.3% | 2.2% | 15.8% | 1.5% | -2.5% | 0.348 | 0.932 |
| m1154 | 7.3% | 2.6% | 12.4% | 1.7% | -5.1% | 0.123 | 2.640 |
| m1136 | 3.3% | 1.5% | 4.9% | 1.0% | -1.6% | 0.396 | 0.759 |
| m1131 | 14.0% | 3.1% | 14.5% | 2.1% | -0.5% | 0.888 | 0.020 |
| m1125 | 23.8% | 3.0% | 28.5% | 2.0% | -4.7% | 0.208 | 1.717 |
| **m1121** | **13.7%** | **1.7%** | **18.3%** | **1.2%** | **-4.6%** | **0.041** | **4.879** |
| m1119 | 8.8% | 2.0% | 10.8% | 1.3% | -2.0% | 0.414 | 0.702 |
| m1107 | 3.2% | 1.7% | 5.4% | 1.1% | -2.2% | 0.287 | 1.209 |
| m1100 | 18.0% | 2.4% | 23.6% | 1.7% | -5.6% | 0.075 | 3.601 |
| m1097 | 21.0% | 2.8% | 26.3% | 1.9% | -5.3% | 0.136 | 2.455 |

**Figure 2:**
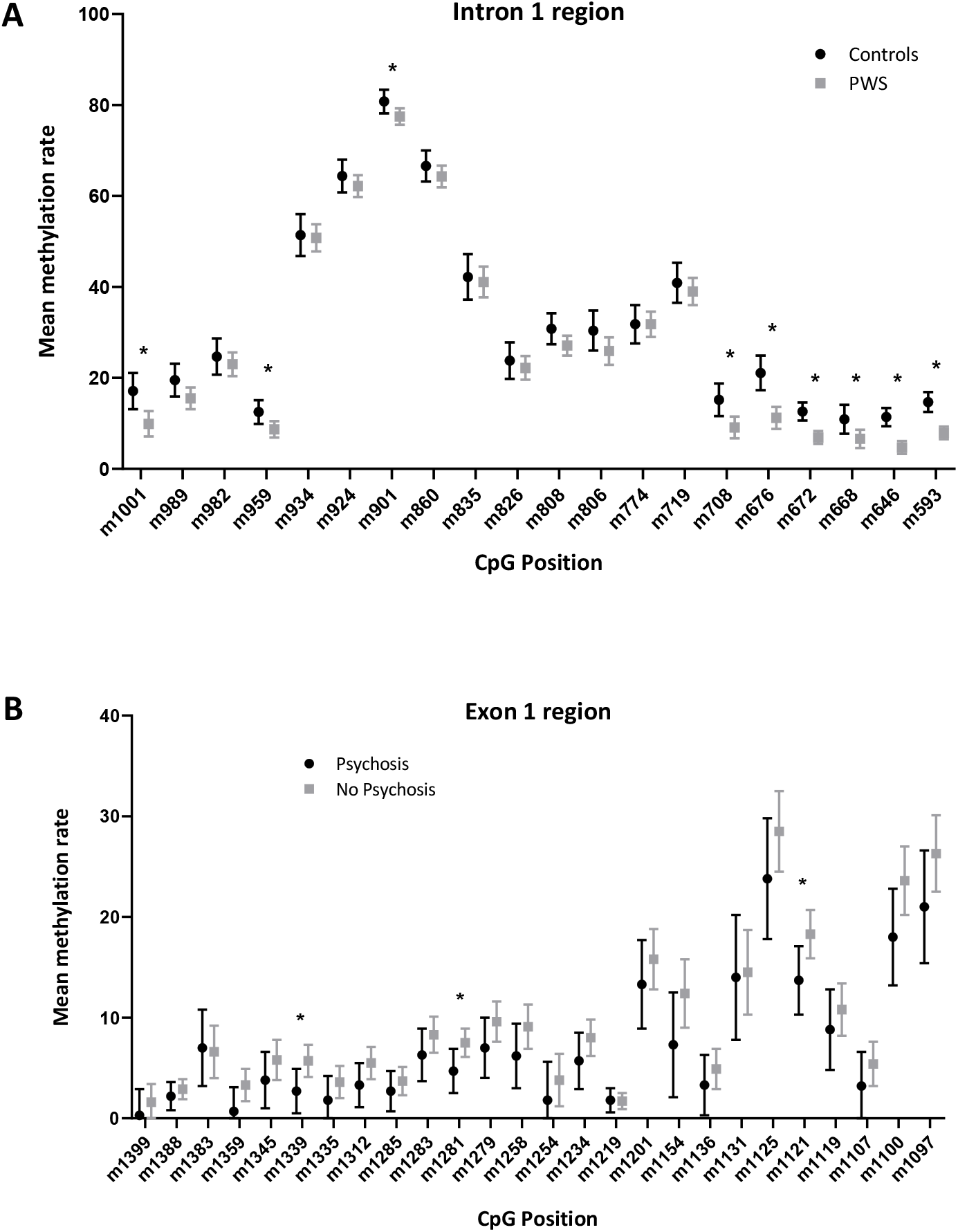
<u>A: Mean methylation rates of PWS subjects compared to controls in the intron 1 region.</u> CpGs where significantly lower methylation rates were detected in PWS subjects compared to controls are indicated by an asterisk. The graph shows mean values and SEMx2 values. <u>B: Mean methylation rates of PWS males with and without history of psychosis in the exon 1 region.</u> CpGs where significantly lower methylation rates were detected in PWS males with psychosis compared to those without psychosis are indicated by an asterisk. The graph shows mean values and SEMx2 values.

#### No significant effects of subtype or psychosis

As the neuropsychiatric phenotype of PWS is quite different between the genetic subtypes of PWS, we also analyzed differences in mean methylation between the genetic subtypes of PWS. We compared the mean methylation rates of the individuals with a deletion subtype with those of the individuals with a mUPD or IC subtype and found no significant differences between these subtypes (F_(1,618)_=0.001, p=0.971). PWS individuals with the deletion subtype showed a mean methylation rate of 27.3% ± 1.3% compared to a mean methylation rate of mUPD and IC subtypes of 27.2% ± 1.2%.

In the next step, we wondered if the status of psychosis (yes/no) affects the mean methylation rates. For the intron 1 region, no significant effect of psychosis on the mean methylation rate was detected (F_(1,618)_=0.081; p=0.776).

#### Analysis of transcription factor binding sites

To asses functional relevance of those CpG sites where a significant difference was found between PWS subjects and neurotypical controls, we checked for possible binding of transcription factors at these sites. The Factorbook ChipSeq database predicted possible binding of transcription factors to each of those CpG sites (table 3). Of particular interest, CpG m1001 was identified as a possible binding site for the transcriptional repressor CTCF, CpG m672 as a possible binding site for the zinc finger protein 142 (ZNF142) and CpG m646 as a possible binding site for the zinc finger and BTB-domain-containing protein 7a.

**Table 3:**
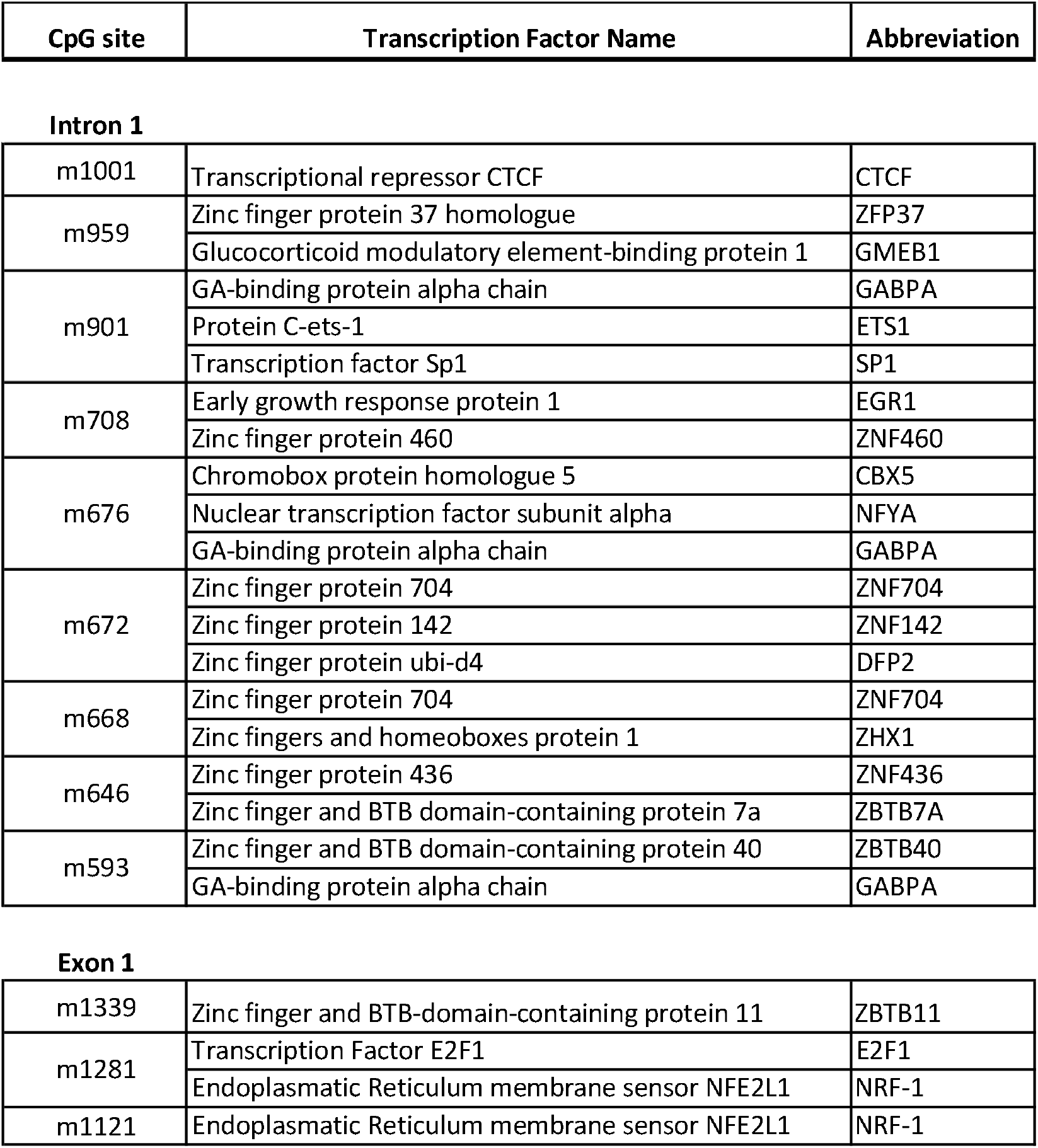
Selection of transcription factors that could possibly bind to CpG sites where methylation differed significantly between the PWS group and the neurotypical controls (intron 1) or between PWS males with a history of psychosis and without history of psychosis (exon 1). Transcription factors listed here were selected from the first 9 results shown by the FACTORBOOK ChipSeq database. The main selection criterion was the functional relevance of the CpG in question for the binding of the query OXTR sequence with the transcription factor sequence (as shown by size of the nucleotide in the motif given by FACTORBOOK). Names of the transcription factors given in this table correspond to the names recommended by Uniprot (https://www.uniprot.org/).

| CpG site | Transcription Factor Name | Abbreviation |
| --- | --- | --- |
| Intron 1 |  |  |
| m1001 | Transcriptional repressor CTCF | CTCF |
| m959 | Zinc finger protein 37 homologue | ZFP37 |
|  | Glucocorticoid modulatory element-binding protein 1 | GMEB1 |
| m901 | GA-binding protein alpha chain | GABPA |
|  | Protein C-ets-1 | ETS1 |
|  | Transcription factor Sp1 | SP1 |
| m708 | Early growth response protein 1 | EGR1 |
|  | Zinc finger protein 460 | ZNF460 |
| m676 | Chromobox protein homologue 5 | CBX5 |
|  | Nuclear transcription factor subunit alpha | NFYA |
|  | GA-binding protein alpha chain | GABPA |
| m672 | Zinc finger protein 704 | ZNF704 |
|  | Zinc finger protein 142 | ZNF142 |
|  | Zinc finger protein ubi-d4 | DFP2 |
| m668 | Zinc finger protein 704 | ZNF704 |
|  | Zinc fingers and homeoboxes protein 1 | ZHX1 |
| m646 | Zinc finger protein 436 | ZNF436 |
|  | Zinc finger and BTB domain-containing protein 7a | ZBTB7A |
| m593 | Zinc finger and BTB domain-containing protein 40 | ZBTB40 |
|  | GA-binding protein alpha chain | GABPA |
| Exon 1 |  |  |
| m1339 | Zinc finger and BTB-domain-containing protein 11 | ZBTB11 |
| m1281 | Transcription Factor E2F1 | E2F1 |
|  | Endoplasmatic Reticulum membrane sensor NFE2L1 | NRF-1 |
| m1121 | Endoplasmatic Reticulum membrane sensor NFE2L1 | NRF-1 |

### Exon 1 region

In the exon 1 region, we found a significant effect of sex in methylation rates (F_(1,1139)_=4.145, p=0.042). Males showed a mean methylation rate of 8.5% ± 0.3% and females showed a mean methylation rate of 7.4% ± 0.4%. Thus, we decided to perform the subsequent analysis separately for male and female subjects.

We then compared mean methylation rates of PWS subjects and neurotypical controls. No significant effect of group could be detected neither in male (F_(1,724)_=0.863, p=0.353) nor in female subjects (F_(1,414)_=0.017, p=0.897). Male controls showed a mean methylation rate of 8.1% ± 0.6% and male PWS individuals showed a mean methylation rate of 8.7% ± 0.4%. The difference between these groups was not significant (F_(1,724)_=0.863, p=0.353). Female controls showed a mean methylation rate of 7.4% ± 0.8% compared to a mean methylation rate of 7.5% ± 0.4% in female PWS individuals. Again, the difference between the two groups was not significant (F_(1,414)_=0.017, p=0.897).

Furthermore, we compared mean methylation rates of PWS individuals with a deletion subtype with those of PWS individuals with a mUPD or IC subtype in male PWS individuals and in female PWS individuals in the exon 1 region, respectively. No significant differences in methylation rates between the deletion subtype and mUPD/IC could be seen neither in males (F_(1,492)_=1,376, p=0.241) nor in females (F_(1,310)_=1,090, p=0.297) subjects. In male PWS subjects, individuals with the deletion subtype showed a mean methylation rate of 9.2% ± 0.5% and individuals with mUPD or IC subtype showed a mean methylation rate of 8.3% ± 0.5%. In female PWS subjects, individuals with a deletion subtype showed a mean methylation rate of 8.1% ± 0.7% and individuals with mUPD or IC subtype showed a mean methylation rate of 7.3% ± 0.5%.

#### Lower methylation in PWS males with psychosis

Again, we wanted to examine if the status of psychosis (yes/no) affects the mean methylation rates. As there was only one female patient with PWS and psychosis we did not perform that analysis in the female group because of a lack of statistical power. In male PWS subjects, significant effects were detected for psychosis on the mean methylation rate (F_(1,492)_=9.325; p=0.002). Male PWS subjects with a history of psychosis showed a lower mean methylation rate of 7.1% ± 0.7% than male PWS subjects without a history of psychosis, who showed a mean methylation rate of 9.5 ± 0.4%. Analysis of methylation rates at single CpG sites showed significant effects of psychosis at CpG site m1339 (F_(1,17)_=4.764; p=0.043), m1281 (F_(1,17)_=4.522; p=0.048) and m1121 (F_(1,17)_=4.879; p=0.041) (table 2B, figure 2B).

#### Analysis of transcription factor binding sites

Analysis of those CpGs as possible binding sites for transcription factors using the open-source TransFac database showed CpG m1281 to be a possible binding site for the Transcription factor E2F1. The probability of binding and the functional relevance of CpG m1281 for the binding of E2F1 to the according OXTR DNA sequence could be confirmed using the position frequency matrices of the open access ChipSeq databases Factorbook and JASPAR. The Factorbook database also showed CpG m1339 to be a possible relevant binding site for the transcription factor zinc finger and BTB domain-containing protein 11 (ZBTB11) (table 3).

## Discussion

In the present study we show that the methylation pattern of the CpG island spanning exon 1 and intron 1 of the OXTR gene is altered in PWS. Several studies indicate an impairment of the oxytocin system in PWS in a sense of a lower oxytocin expression. For instance, a decrease of OT-expressing neurons was found in a *post mortem* study of PWS patients [14]. This finding was supported by a more recent study, where both the OT mRNA levels and the number of cells immunoreactive for OT were reduced in the PVN of individuals with PWS [36]. Furthermore, in PWS murine models, a knockout of Necdin and MAGEL2 led to a reduction in OT-producing neurons [17, 18]. Höybye et al. found peripheral OT levels to be within the normal range, but abnormally low when taking the obesity of PWS patients into account [15]. Contrarily, another study reported elevated OT levels in children with PWS compared to healthy siblings matched by gender, age and BMI [16]. Nonetheless, plasma OT levels do not necessarily reflect central OT levels, as OT is secreted both centrally via axon terminals and peripherally into the bloodstream via the neurohypophysis [37]. Our results of a significant hypomethylation of CpG sites located in the intron 1 of the OT receptor gene (*OXTR*) in PWS subjects compared to neurotypical controls indicate an alteration of the OT system in PWS also on an epigenetic level. The region of the *OXTR* investigated in this study covers a region previously described by Kusui et al. to regulate expression of the OT receptor, with higher methylation rates indicating reduced transcription of the oxytocin receptor [38]. A recent study confirmed that DNA methylation in this region negatively correlates with the expression of the OT receptor in human temporal cortex [39]. Consequently, the hypomethylation we observed indicates an increased expression of the OT receptor in PWS individuals, which might act as a potential compensatory mechanism of the decreased OT production seen in PWS. In contrast to this, a whole genome microarray analysis of gene expression and RT-PCRs in PWS males revealed a reduction in the expression of the OT receptor in both lymphoblastoid cells and frontal cortex tissue of PWS individuals compared to control males [40]. However, RNA is known to be unstable, which specifically makes it hard to accurately examine expression levels in *post mortem* tissue.

Individuals with PWS have a higher susceptibility for developing psychosis than the general population, which is particularly evident for the mUPD subtype with a prevalence of up to 62% of psychosis [41]. Regarding psychotic disorders in general, an implication of the OT system has been described previously as well. Although normal peripheral OT levels have been reported in psychotic patients, several SNPs in the *OXTR* gene are associated with psychosis, supporting the hypothesis that the OT system is affected at the receptor level (Davis et al., 2014; Montag et al., 2013; Rubin et al., 2016). A previous study showed higher methylation of a single CpG site within the intron 1 region of the *OXTR* in patients with prototypic schizophrenic features than in controls [30]. In our study, we found the CpG sites spanning exon 1 to be significantly hypomethylated in PWS patients with a history of psychotic episodes compared to those without, which seems contrary to the findings of Rubin et al. However, psychosis in PWS rather has affective symptoms similar to psychotic episodes found in bipolar disorders as opposed to typical schizophrenic symptoms [42, 43]. Interestingly, the above mentioned study found methylation in patients with psychotic bipolar disorder to be lower than in patients with prototypic schizophrenia [30], which supports our findings.

As we measured the methylation levels using peripheral blood, one could argue that they do not reflect central activity. However, evidence suggests that for the *OXTR*, peripheral blood DNA methylation status can be used as a biomarker for the methylation and transcription status of the *OXTR* in brain. For instance, peripheral DNA methylation levels have predicted central levels (nucleus accumbens) in prairie voles [44]. Furthermore, a positive association between peripheral blood and temporal cortex *OXTR* methylation has been found in human ASD individuals [23].

Methylation of even single CpG sites might lead to changes in expression by interfering with transcription factors (TF) that would normally bind to the unmethylated CpG single sites [45, 46]. We found numerous TF that likely bind at CpG sites where methylation significantly differed between individuals with psychosis and no psychosis and PWS subjects in general and neurotypical controls. Among these TF were several that have previously been associated with neurodevelopmental disorders: Firstly, the Transcriptional repressor CTCF, which can bind to CpG m1001, has been shown to play a role in neurodevelopmental processes and has been linked to characteristics also seen in the PWS phenotype such as intellectual disability, behavioral challenges, feeding difficulties and growth restriction [47–49]. Furthermore, we identified CpG m672 as a possible binding site for ZNF142, CpG m1339 as a possible binding site for ZBTB11 and CpG m646 as a possible binding site for ZBTB7a. These TF are associated with intellectual disability and might as well be involved in nervous system development [50–54].

In our opinion, another TF is especially noteworthy: Transcription factor E2F1, also called Retinoblastoma-associated protein 1 (RBAP1), likely binds at CpG m1281, which was hypomethylated in male PWS subjects with a history of psychosis. E2F1 mediates apoptosis of neurons, plays a key role in regulating neuronal differentiation and promotes proliferation of neural progenitors in the developing mammalian hypothalamus [55–57]. Intriguingly, E2F1-dependent transcription has been shown to be repressed by the anti-apoptotic protein Necdin, a gene located within the PWS locus on Chromosome 15q11-q13 and thus not expressed in PWS [58–60]. Necdin has been shown to suppress both proliferation and apoptosis of neocortical neural stem/progenitor cells (NSPCs) by binding to the C-terminal transactivation domain of E2F1 and thereby suppressing E2F1-dependent transactivation of Cyclin-dependent kinase 1 (Cdk1) [56, 58]. The hypomethylation in PWS males with psychosis seen in our study could thus reflect a dysregulation of proliferation and apoptosis during neurodevelopment caused by lack of Necdin-mediated inhibition of E2F1. Interestingly, as mentioned above, Necdin-deficient mice also display fewer OT-producing neurons [18].

The finding of altered expression levels in numerous receptor genes (e.g. GABA, serotonin and oxytocin receptor genes) in PWS has sparked the thesis of a common pattern of transcriptional regulation of those receptor genes that might be regulated by genes on Chr. 15q11-q13 [40]. The identification of a TF regulated by Necdin in a receptor gene with a differential methylation pattern in PWS subjects in our study supports this hypothesis. Furthermore, it seems that this proposed mechanism could also contribute to the development of psychiatric disorders, such as psychosis, in PWS.

## Limitations

There are several limitations to our study. We are aware that the sample size of our study, especially the sample size of PWS individuals with psychosis, is relatively small. However, taking the rarity of PWS into account, our cohort is one of the largest investigating epigenetic changes in PWS regarding the available literature so far. Furthermore, we did not investigate group differences in PWS individuals with and without ASD as we did not include diagnosis of ASD in our study, even though atypical methylation of the *OXTR* has previously been shown in ASD subjects [23, 26, 61]. In this study, we focused on the methylation status in the exon 1 and intron 1 region. Further studies could include measurements of methylation status in the exon 3 and intron 3 region as well, as several SNPs and different methylation patterns in those regions have been associated with psychiatric disorders such as ASD and schizophrenia and with psychosocial stressors in healthy individuals [19–21, 25, 62–65].

## Conclusion

Taking together, we are the first to identify *OXTR* CpG sites that are significantly dysregulated in PWS subjects and individuals with PWS and psychosis. We conclude that two different processes might influence the methylation pattern of the OXTR in PWS. One, a decreased production of OT in PWS might lead to a compensatory increased expression of OT receptors by means of DNA hypomethylation. Additionally, a possible differential transcriptional regulation of receptor genes in PWS might interfere with the development of a physiological DNA methylation pattern in the *OXTR* promoter region and thereby could contribute to the development of psychosis in PWS.

## Data Availability

The datasets generated and/or analysed during the current study are not publicly available due to data privacy reasons but are available from the corresponding author on reasonable request.

